# Weed Out the Risk: Pharmacovigilance in Medical Cannabis Users

**DOI:** 10.1101/2025.07.18.25331800

**Authors:** Mitchell L. Doucette, Junella Chin, Emily Fisher

## Abstract

**Introduction:** Medical cannabis use has expanded rapidly, yet long-term real-world safety data remain limited. We evaluated adverse-event (AE) frequency, severity, and predictors in a US telehealth registry of medical cannabis patients over one year.

**Methods:** We analyzed 14,313 adults who completed intake between June–August 2024. Patients reported any of 30 prespecified adverse events (AEs) and rated each on a 0–10 impact scale. Weekly exposure was estimated as (days/week) × (serving size) and categorized into quintiles. We computed AE rates per 100 patients with binomial 95% confidence intervals (CI) and tested linear trends. Univariate logistic regressions assessed 20 candidate predictors within chronic-pain and anxiety subgroups. We then applied LASSO to select multivariate predictors, combining these with age, sex, race/ethnicity, smoking, and unhealthy-weeks in final logistic models. Marginal predicted-probability curves were generated across exposure, stratified by subgroup, sex, race, and age.

**Results:** Overall, 2.6% of patients reported ≥1 AE. The most common symptoms were increased appetite (23.8%), fatigue (20.3%), and anxiety (19.9%) with mean impact <4/10. In adjusted models, having been to the doctor because of their condition remained the sole AE predictor for patients with anxiety (OR 4.03, 95% CI: 2.44–6.87); age was a significant predictor for patients with chronic pain (OR 0.981, 95% CI: 0.97–0.99). Marginal curves remained flat (∼2–3% AE probability) across weekly cannabis exposure. Ad hoc analysis of non-missing-at-random data suggests possible AE rates are in line with current literature.

**Discussion:** In this large cohort, AEs were infrequent and mild, and weekly cannabis frequency did not independently increase odds. Healthcare engagement likely reflects underlying health complexity driving AE reporting. These findings support the safety of medical cannabis.

## 1 Introduction

The use of medical cannabis as a therapeutic has increased greatly in the U.S. and abroad over the past decade (Boehnke et al., 2024; Wang et al., 2024). Indeed, the 21^st^ century has seen cannabis legalized for medical purposes in the UK, Canada, Australia, Germany, Israel, a majority of the US, and several other nations (Schlag, 2020; Shover and Humphreys, 2019). The increased ubiquity of the medical product has shifted public opinion positively towards the concept of cannabis as medicine (CaM) (Hallinan et al., 2022; Hossain and Chae, 2024); Descriptive trend analyses show increases in overall medical cannabis use (Boehnke et al., 2024; Wang et al., 2024).

Cannabis is one of the oldest medicinal plants in the world containing over 500 reported compounds, and specifically 125 isolated cannabinoids (Lu and Mackie, 2016; Radwan et al., 2021). The plant contains two primary constituents, cannabidiol (CBD) and delta-9-tetrahydrocannabinol (THC) with many other cannabinoids, such as tetrahydrocannabivarin (THCV), being identified as potentially therapeutic. These cannabinoids interact with the endocannabinoid system (ECS), an important neuromodulator system that helps maintain physiological balance by producing lipid-based messengers (endocannabinoids) on demand, which bind to CB1 and CB2 receptors to broadly tune neuronal and non-neuronal signaling. In doing so, the ECS regulates processes such as pain perception, appetite, mood, memory and immune function (Lu and Mackie, 2016).

Recent research has demonstrated that CaM can prove efficacious for a number of health conditions. The seminal 2017 National Academies of Science reported strong evidence for the use of medical cannabis in the treatment of chronic pain, multiple sclerosis spasticity, and as an antiemetic (National Academies of Sciences, Engineering, and Medicine, 2017). Newer research has found some evidence of efficacy specific to post-traumatic stress disorder (Doucette et al., 2024d; Krediet et al., 2020; Lynskey et al., 2024; Nacasch et al., 2023; Pillai et al., 2022; Rehman et al., 2021; Roitman et al., 2014; Sznitman et al., 2020), sleep disorders (Ried et al., 2023; Sznitman et al., 2020; Vivek et al., 2024; Walsh et al., 2021), and inflammatory bowel disease (Dalavaye et al., 2023; Greywoode et al., 2023; Sunil et al., 2022) among other medical conditions. Other research has found medical cannabis to assist with discontinuation of potentially harmful medications such as prescription opioids (Jeddi et al., 2024; Nguyen et al., 2023; O’Connell et al., 2019; Shi, 2017; Takakuwa et al., 2020; Wen and Hockenberry, 2018) and benzodiazepines (Doucette et al., 2025b; O’Connell et al., 2019; Purcell et al., 2019), and produce downstream effects on healthcare utilization (Cook et al., 2023; Doucette et al., 2024c; Greywoode et al., 2023; Sobotka et al., 2021).

As legalization and public demand for CaM have expanded, research into its potential benefits and risks has likewise intensified and its use is not without safety concerns (MacCallum and Russo, 2018; MacCallum et al., 2021). Several potential adverse events (AEs) of medical cannabis have been identified with increased research. Most of the AEs are associated with THC rather than CBD, and are related to psychoactivity, specifically dizziness, psychological disturbances, and somnolence (Gottschling et al., 2020). Though other common AEs exist, such as dry mouth, fatigue, nausea, vomiting, headache, constipation, and muscular weakness. Researchers have increasingly completed controlled studies with real-world data to generate pragmatic evidence on CaM safety (Crescioli et al., 2020; Dalavaye et al., 2023; Ergisi et al., 2022; Erridge et al., 2022, 2023; Hachem et al., 2024; Harris et al., 2022; Kalaba et al., 2022; Nicholas et al., 2023; Nimalan et al., 2022; Olsson et al., 2023; Pillai et al., 2022; Rifkin-Zybutz et al., 2023; Tait et al., 2023; Vivek et al., 2024).

### 1.1 Safety Evidence from Clinical Trials and Real-World Evidence

Across multiple RCTs, including crossover and parallel-group designs, pharmacovigilance data reveal low rates of treatment-related AEs (Abrams et al., 2020; AminiLari et al., 2022; Aran et al., 2021; Grimison et al., 2024; Zylla et al., 2021). For example, in adults with sickle-cell disease, inhaled cannabis (treatment group) produced a similar discontinuation rate as the placebo group and with no serious or severe events attributable to treatment (Abrams et al., 2020). In a proof-of-concept autism trial, whole-plant and pure CBD preparations yielded only mild, non-serious AEs (Aran et al., 2021). While the whole-plant and CBD groups had higher a percentage of patients report somnolence as an AE compared to placebo, all other common AEs occurred at similar percentages across the three trial arms (Aran et al., 2021). A Phase II/III RCT of oral extract for chemotherapy-induced nausea documented increased sedation in 18% and increased dizziness in 10% of treated patients versus 7% or less in placebo, with only two discontinuations due to neuropsychiatric effects (2.8% discontinuation rate) (Grimison et al., 2024). Other common AEs, specifically anxiety and disorientation, were not statistically significantly different comparing treatment to placebo groups (Grimison et al., 2024). A meta-analysis of 39 sleep-specific RCTs found medical cannabis was associated with small risk differences (RD) in the following AEs, nausea, vomiting, headache, fatigue, dry mouth, dizziness, nausea, and somnolence. However, overall evidence suggested medical cannabis was associated with small benefits in sleep disturbances for those with impaired sleep (AminiLari et al., 2022). A randomized trial of medical cannabis patients with stage IV cancer (n = 30) found no serious safety issues and high satisfaction from patients who were randomized to either early or delayed start cannabis (Zylla et al., 2021).

Real-world data from observational registries extend safety surveillance to broader, more heterogeneous populations. Results from the UK’s medical cannabis database have documented safety signals across a large swath of medical conditions including anxiety, chronic pain, and PTSD, among others (Dalavaye et al., 2023; Ergisi et al., 2022; Erridge et al., 2022, 2023; Harris et al., 2022; Nicholas et al., 2023; Nimalan et al., 2022; Olsson et al., 2023; Pillai et al., 2022; Rifkin-Zybutz et al., 2023; Tait et al., 2023; Vivek et al., 2024). While most of these registry-based examinations contain small populations, they provided baseline and follow-up information, including safety signals. The AE rate across all 11 manuscripts published using the UKs medical cannabis patient registry vary, from 47% in the trial of children treated for treatment resistant epilepsy to 13.1% for adults with insomnia. In all of the condition-specific examinations, the average AE reported was greater than 1 per person and most were non-serious.

Other countries with centralized data related to medical cannabis use report similar AE rates to the UK medical cannabis database and provided descriptive information on AE demographics. The Italian Phytovigilance database (252 patients) recorded an adverse reaction rate of 21%, though they were also primarily non-serious (Crescioli et al., 2020). The mean age was 61.9 and was primary female (77.3%). Nearly 99% of all AE came from an oral cannabis administration route, compared to an inhalation route, with nearly 70% of AE completely resolving. Quebec’s pharmacovigilance registry (2,991 participants) reported moderate or severe AEs in just 3.6% of patients, prompting discontinuation in fewer than 0.3% (Hachem et al., 2024). The mean age of those with an AE was 50.9 years and females reported 50.2% of AEs. An Australian real-world data study (535 participants) noted that 57.9% of patients reported at least one AE, 97% of which were non-serious with a discontinuation rate of 6% (Kalaba et al., 2022).

High-level evidence syntheses have consolidated cannabis’ safety signals across diverse study designs. A scoping review of 72 systematic reviews found minor AEs such as dizziness, dry mouth, and nausea in more than half of analyses. Serious harm was not common, but at least one serious harm was reported in 36% of the 72 reviews, often without definitive causality attribution (Pratt et al., 2019). An umbrella review of 101 meta-analyses reported elevated odds ratios for central nervous system events (OR 2.8), psychological effects (OR 3.1), and visual disturbances (OR 3.0), as well as improved nausea/vomiting, pain, and spasticity (Solmi et al., 2023). An earlier systematic review of 31 studies documented that 96.6% of all AEs were non-serious and 3.4% were serious, including relapse of multiple sclerosis and gastrointestinal complications (Wang et al., 2008).

### 1.2 Current Contribution

Despite an expanding body of trial and registry data, long-term safety signals for medical cannabis remain characterized by small cohorts and brief follow-up. To add to this body of literature, we conducted a pharmacovigilance study using a US-based real-world data registry of over 14,000 patients who used medical cannabis for 1 year. As such, we had the following three aims: 1) describe the AE frequency and rate per 100 patients, as well as the self-reported tolerability of the AEs, across demographics, lifestyle characteristics, and cannabis use habits, 2) identify predictors of AEs across top two conditions (anxiety and chronic pain), and 3) examine whether the probability of having an AE varies based on weekly cannabis exposure across select demographics and conditions.

## 2 Methods

### 2.1 Study Design and Population

We conducted a retrospective pharmacovigilance study using real-world data from a registry of patients seeking medical cannabis certification through Leafwell, a national telehealth platform. This database has been previously used for academic purposes (Doucette et al., 2024d,b,a, 2025a,b). Leafwell is a telehealth platform that facilitates access to licensed physicians for patients who may qualify for medical cannabis within a given state. The company operates in 34 states and advertises on internet search engines and digital media. As part of being seen by a qualified physician, patients must fill out an intake form regarding demographic, lifestyle behaviors, and for those renewing their certification cards, past-year medical cannabis use habits. We utilized the data from the intake form to inform our study.

We accessed administrative data from June 2024 to August 2024 (3-month study period) for returning patients to Leafwell, or those individuals who had previously been certified to use medical cannabis 1 year earlier and came back to become re-certified during the study period. For these individuals, their intake form asked them to self-report medical cannabis use habits for the past year and any AEs. We excluded any patients under the age of 18. Internal researchers did not have access to non-anonymized data, and this project received exempt status from an external third-party institutional review board (IRB).

### 2.2 Measures

In the Leafwell intake form, patients were asked whether they experienced any cannabis-related AEs in the past year (yes vs. no). If they responded positively, patients were asked to identify which AEs they had suffered from a list of 29 specific AE, including an ‘Other’ option (Full list provided below). For each AE that was selected, there was a corresponding question to assess its severity; patients were asked to rate, on a scale of 0 to 10, with ‘0’ being No impact at all and ‘10’ Extremely impactfull, how the AE impacted their daily life.

We included several other covariates in this analysis. Available demographic data included age (continuous), sex (male vs. female), race/ethnicity (non-Hispanic, white vs. all other races), education (college degree or higher vs. less than a college degree), veteran status (yes vs. no) and health insurance status (yes vs. no). We also included data related to patient’s tobacco smoking status (current smoker vs. former or never smoker) alcohol consumption habits (non-drinkers vs. those who drink at least one day per week).

We included two validated measures of health status. First, we used the Centers for Disease Control’s Health-related Quality of Life-4 scale, which provided a number of unhealthy days per month (Mielenz et al., 2006; Zahran et al., 2005). We created a four-level variable representing the unhealthy weeks per month (1 week or less, 2 weeks or less, 3 weeks or less, and more than 3 weeks per month). We also used the Graded Chronic Pain Scale Revised to assess pain severity within the study population (Von Korff et al., 2020). We created a binary variable comparing no/mild chronic pain to bothersome/severe chronic pain.

Patients were asked a set of cannabis use habit questions. Patients were asked to report how many days per week they used medical cannabis (1 through 7 days), their most common route of administration (dried flower (vaporizer), dried flower (smoked), edible, inhaler, sublingual spray, vape pen, tablet/capsule, tincture/oils/drops, topical lotion, or transdermal patch), and the type of product they most commonly used (High THC vs Low CBD, Low THC vs High CBD, THC only, CBD only, Equal amounts of CBD and THC). For each route of administration, patients were asked to specify the most common serving size, which ranged from less than 1 serving to 4 or more servings (Supplemental Table 1 provides serving size information).

#### 2.2.1 Weekly Cannabis Exposure

To assess dose-response relationships, we constructed a weekly cannabis exposure variable. To accomplish this, a composite continuous variable for weekly exposure was created by multiplying the number of days per week a patient consumed by the average serving size per use. Serving size estimates were harmonized across modalities (see Supplemental Table 1 for more detail). For example, 1 edible = 1 smoked bowl (dried flower). This variable was used as a continuous predictor. We also presented this variable in quintiles to examine whether the risk of AEs increased as the measure of weekly cannabis exposure increased.

### 2.3 Statistical Analysis

All analyses were conducted in R (version 4.5.1) (RStudio Team, 2025). We first generated descriptive statistics: counts and percentages (for categorical variables) and means ± SD (for continuous variables) were compared between patients reporting any AE and those reporting none, using *χ*^2^ tests for categorical data and two-sample t-tests for continuous variables. We also calculated unadjusted AE rates per 100 patients with exact binomial 95% confidence intervals (CIs).

Next, we characterized the AE experience itself. First, we examined AE prevalence across weekly-exposure quintiles, computing within-quintile counts, proportions, binomial 95% CIs, and testing for a linear trend in the log-odds of AE using a Cochran–Armitage approach (Cochran, 1954; Armitage, 1955; Signorell et al., 2025). We then tabulated the number and proportion of patients who reported each of the pre-specified AEs, and summarized mean impact scores (0–10 scale) with standard errors. We did that overall and stratified by patients with anxiety and patients with chronic pain.

Given the small prevalence of AEs within the sample, we used a multi-step process to identify meaningful predictors of AEs within the two largest patient subgroups, chronic pain and anxiety. First, we performed univariate logistic regressions with each covariate entered singly into a logistic model with AE (yes vs no) as the outcome; coefficients, 95% CIs, and p-values were extracted. We applied robust “sandwich” standard errors to all regression models to mitigate potential misspecification of the binomial variance. Missingness was minimal (*>* 90% complete for all covariates).

We then sought to identify the most relevant predictors of having an AE to ensure the most parsimonious model. To this end, we next fitted penalized logistic models (LASSO, *α* = 1) with 5-fold cross-validation using the R package, glmnet (Tibshirani et al., 2010; Friedman et al., 2008). The design matrix included weekly exposure and all other covariates. We selected non-zero coefficients at the *λ* that minimized cross-validated deviance (*λ* min), yielding a small set of LASSO-selected predictors (Tibshirani, 1996). We used these selected coefficients in our final multivariate logistic models to identify predictors of AE in addition to the following a prior variables, sex, race/ethnicity, age, tobacco use status, number of unhealthy weeks, and weekly cannabis exposure. We extracted adjusted odds ratios (ORs) with robust standard errors and 95% CIs.

Finally, we visualized the exposure–response relationship via marginal predicted-probability curves. For each model, we created a 200-point exposure grid spanning the observed weekly-use range, held all other covariates at reference values (mean age; female; non-White; non-smoker; no doctor visit; *<* 1 unhealthy week), and used predict probabilities to obtain logit-scale fits and standard errors. We back-transformed to probabilities and constructed 95% logit-scale CIs. We expressed the marginal exposure-response curves overall for anxiety and chronic pain patients. Within both anxiety and chronic pain patients, we also visualized the exposure-response curve stratified by sex and race/ethnicity. Additionally, we examined the predicted probability of having an AE across the age spectrum.

## 3 Results

A total of 14,313 patients were included in the study. This represented 59.7% of the total patient population that was eligible for medical card renewal during the 3-month study period. Of them, 367 reported having at least one AE, or (2.6%). Of the 367 patients who reported an AE, 261 reported what their AEs was (71.1%). In total, there were 375 reported AEs among the 261 patients who specified what their AEs were, or around 1.44 AEs per patient.

Those who experienced an AE were slightly younger (mean 43.2 vs. 45.1 years; p = 0.013) and more often female (56% vs. 49%; p = 0.009) (Table 1). There was no significant difference by race/ethnicity (White non-Hispanic 79% vs. 75%; p = 0.13) or veteran status (6.5% vs. 6.6%; p *>* 0.9). College graduates had a modestly higher AE rate than those without a degree (2.96 per 100 patients vs. 2.33 per 100 patients). Current smokers and non-smokers reported similar AE frequencies (18% vs. 20%; p = 0.40), as did drinkers versus non-drinkers (35% vs. 65%; p = 0.80).

**Table 1:**
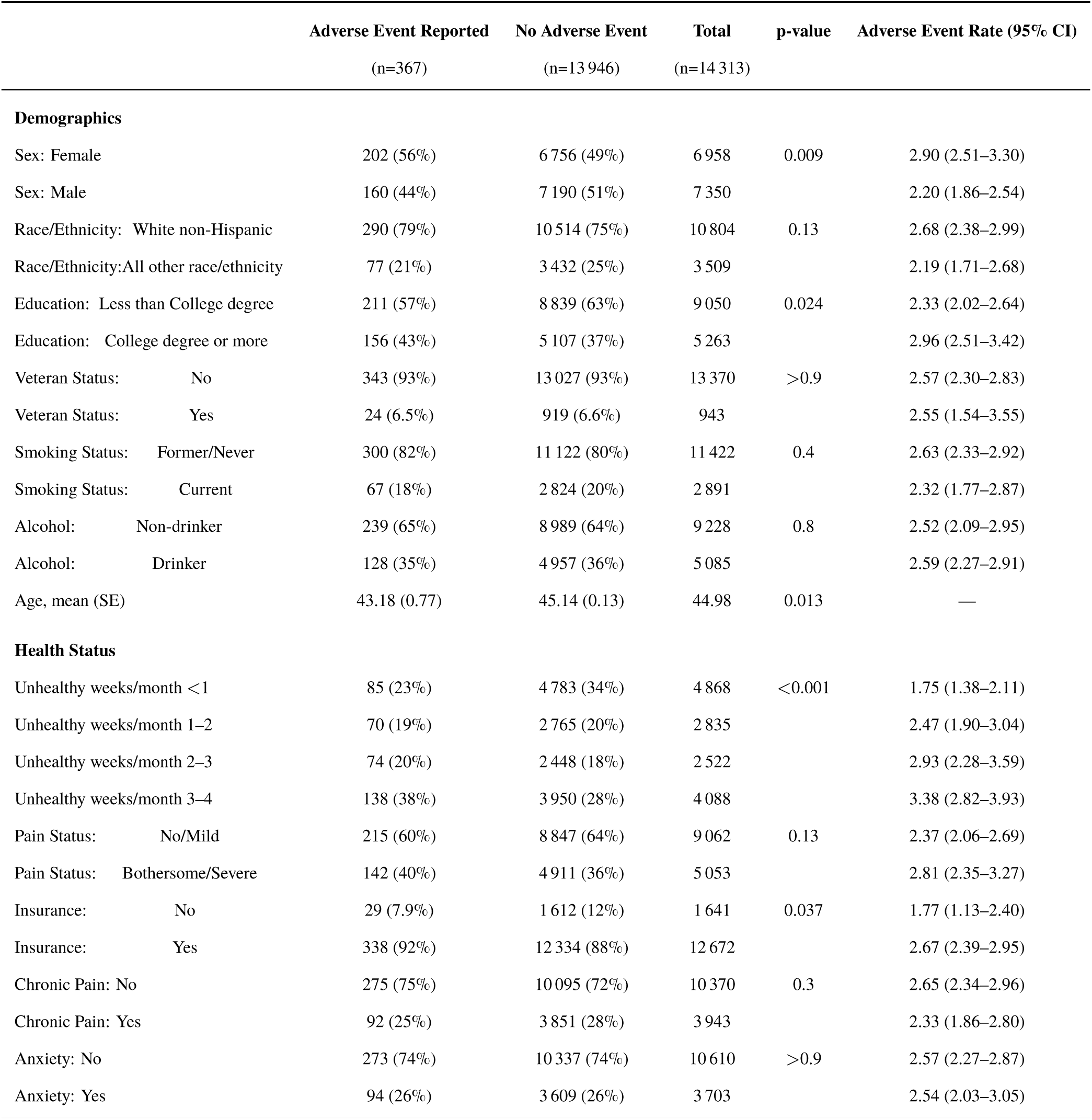
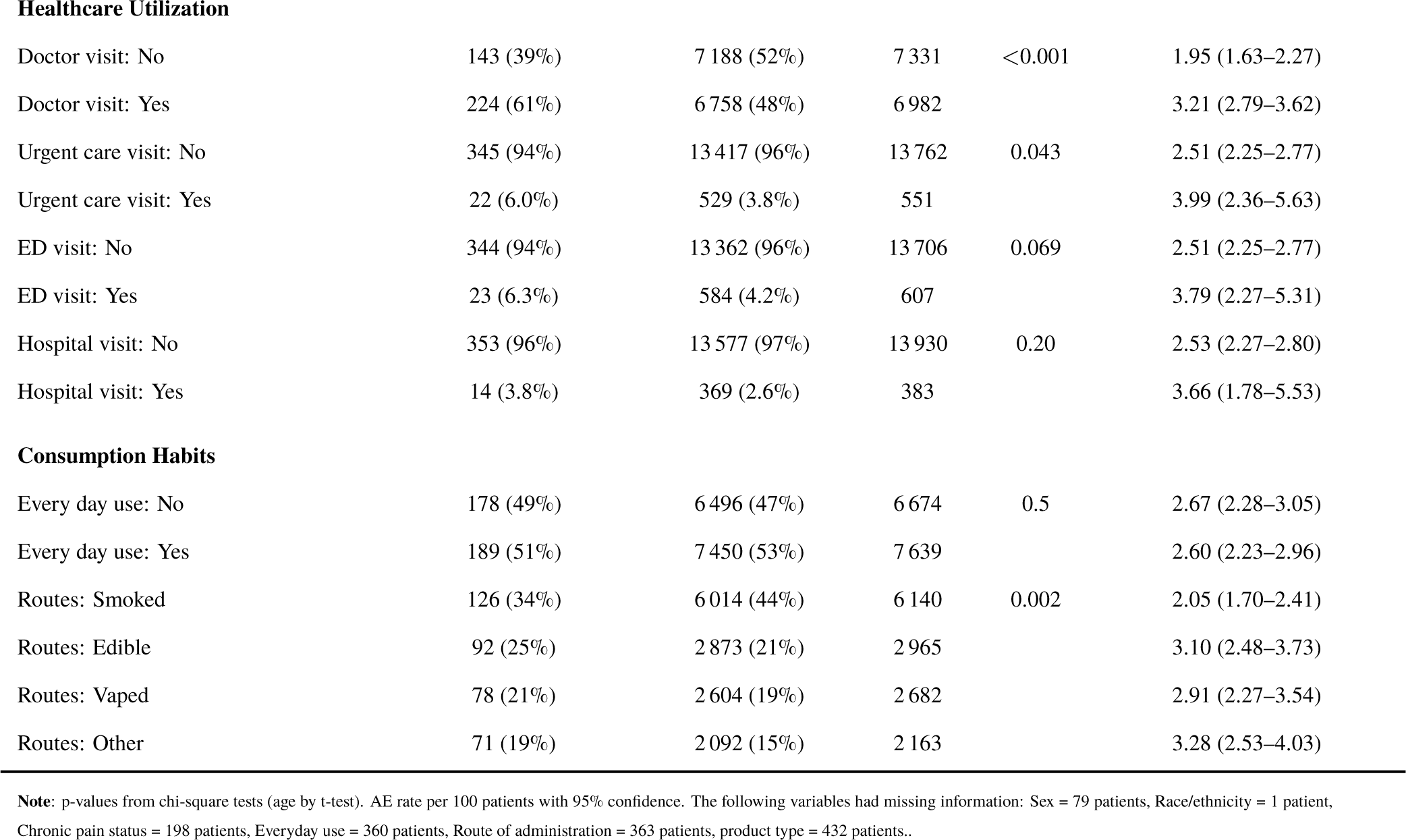
Descriptive table of covariates across the presence of at least one adverse event.

Patients with more unhealthy weeks per month were increasingly likely to report AEs (from 1.8 per 100 in *<*1 week to 3.4 per 100 in 3–4 weeks; p *<* 0.001) (Table 1). Self-reported pain severity and chronic pain or anxiety diagnoses did not significantly affect AE risk. Those who sought healthcare for their condition, particularly doctor visits, had markedly higher AE rates: 3.21 per 100 versus 1.95 per 100 for those who did not see a doctor (p *<* 0.001). Urgent-care visits (3.99 per 100 vs. 2.51; p = 0.043) and emergency-department visits (3.79 per 100 vs. 2.51; p = 0.069) also trended toward higher AE reporting. Hospitalizations were uncommon but associated with a higher AE rate (3.66 per 100 vs. 2.53; p = 0.20). Patterns of cannabis use, daily versus less frequent, route of administration, and product type, showed some variation: edible and “other” routes had higher AE rates (3.10 and 3.28 per 100, respectively) compared with smoking (2.05 per 100; p = 0.002), whereas product type did not significantly influence AE frequency (p = 0.80). In general, most variables were not missing data (see Table 1 for missingness details).

AE prevalence was not uniform across weekly-exposure quintiles (Table 2). In the lowest and second lowest exposure quintile (EQ), 84 and 83 of 2,756 patients each reported an AE. Prevalence dipped in EQ3 (63/2,756; 2.29%, 1.9–2.9%) and fell to its lowest in EQ5 (61/2,755; 2.21%, 1.7–2.8%). The results of the Cochran-Armitage trend test suggested AE prevalence declined with increased weekly exposure (p = 0.036). In total, 3.7% of respondents didn’t answer either the daily use question (n = 360) or the average serving size question (n = 174). Due to the low percentage of data missingness for this exposure variable, we utilized a complete-case analysis.

**Table 2:** Prevalence of Any Adverse Event by Weekly Cannabis Exposure Quintile.

| Exposure Quintile | N | AE Count | Prevalence | 95% CI |
| --- | --- | --- | --- | --- |
| Quintile 1 | 2756 | 84 | 3.05% | (2.4%–3.8%) |
| Quintile 2 | 2756 | 83 | 3.01% | (2.4%–3.7%) |
| Quintile 3 | 2756 | 63 | 2.29% | (1.9%–2.9%) |
| Quintile 4 | 2756 | 73 | 2.65% | (2.1%–3.3%) |
| Quintile 5 | 2755 | 61 | 2.21% | (1.7%–2.8%) |

**Note:** p-value for linear trend in log-odds across quintiles (Cochran–Armitage test) = 0.036. Of the 14,313 patients, 360 (2.5%) did not respond to the question of use habits. An additional 174 people (3.7% total) did not respond to the question related to serving size.

Among the 375 total AE reports captured (Table 3), the most commonly reported symptom was an increase in appetite (n = 62, 23.8%), followed by tiredness/fatigue (n = 53, 20.3%) and anxiety (n = 52, 19.9%) (non-mutually exclusive). These top three AEs also exhibited moderate impact on daily life, with mean impact scores of 3.81 (Standard error (SE) = 0.25) for increased appetite, 3.85 (SE = 0.26) for fatigue, and 3.63 (SE = 0.32) for anxiety. Feeling sick or nauseous and increased cannabis tolerance were each reported by 31 patients (11.9%), with mean impacts of 2.61 (SE = 0.41) and 4.16 (SE = 0.39), respectively.

**Table 3:** Frequency and Mean Impact on Daily Life for Reported Adverse Events.

| Adverse Event | n | % | Mean Impact (SE) |
| --- | --- | --- | --- |
| Increase in appetite | 62 | 23.8 | 3.81 (0.25) |
| Tiredness/fatigue | 53 | 20.3 | 3.85 (0.26) |
| Anxiety | 52 | 19.9 | 3.63 (0.32) |
| Feeling sick or nauseous | 31 | 11.9 | 2.61 (0.41) |
| Increased tolerance to cannabis medication | 31 | 11.9 | 4.16 (0.39) |
| Paranoia | 20 | 7.7 | 3.75 (0.66) |
| Memory loss | 20 | 7.7 | 4.15 (0.35) |
| Weight gain | 18 | 6.9 | 4.39 (0.44) |
| Dizziness | 14 | 5.4 | 3.14 (0.65) |
| Other | 12 | 4.6 | 2.42 (0.50) |
| Sore throat | 9 | 3.4 | 3.33 (0.87) |
| Decreased appetite | 7 | 2.7 | 4.71 (0.84) |
| Diarrhea | 5 | 1.9 | 2.40 (0.40) |
| Loss of balance | 5 | 1.9 | 4.60 (1.63) |
| Irritability | 5 | 1.9 | 4.60 (0.81) |
| Hallucinations (visual and/or auditory) | 4 | 1.5 | 2.50 (0.87) |
| Constipation | 3 | 1.1 | 3.67 (1.33) |
| Negative behavioral or mood change | 3 | 1.1 | 5.33 (0.33) |
| Somnolence | 3 | 1.1 | 3.33 (0.88) |
| Loss of interest in usual activities | 3 | 1.1 | 6.00 (1.00) |
| Agitation | 3 | 1.1 | 4.00 (1.53) |
| Weakness | 2 | 0.8 | 3.00 (2.00) |
| Increased pain sensitivity | 2 | 0.8 | 4.50 (1.50) |
| Wheezing | 2 | 0.8 | 5.00 (1.00) |
| Seizures | 2 | 0.8 | 3.00 (0.00) |
| Increase in depression | 1 | 0.4 | 5.00 (—) |
| Weight loss | 1 | 0.4 | 0.00 (—) |
| Euphoria | 1 | 0.4 | 4.00 (—) |
| Tremors | 1 | 0.4 | 0.00 (—) |

**Note:** Three adverse events (hospitalization, suicidal thoughts, spasms/tics) had zero reports and were excluded. Patients asked to rate, on a scale from 0 to 10, how the adverse event impacted their daily life with 0 being no impact at all and 10 being extremely impactfull. Patients had the ability to report multiple adverse events.

Less frequent events, such as paranoia and memory loss (both 7.7%), showed higher mean impacts (3.75 (SE = 0.66) and 4.15 (SE = 0.35)), while weight gain (6.9%; 4.39 (SE = 0.44)) and dizziness (5.4%; 3.14 (SE = 0.65)) fell in the mid-range of impact. A heterogeneous group of ‘Other’ symptoms (4.6%) averaged 2.42 (SE = 0.50) in impact. Several infrequent AEs, decreased appetite, diarrhea, loss of balance, irritability, had mean impacts from 3.67 to 6.00 but were reported by fewer than 10 patients each (*<* 3%). Rare events such as seizures, wheezing, and increased pain sensitivity (*<* 1% each) demonstrated variable impact (impact range 3–5). Euphoria and weight-loss reports (each 0.4%) were too sparse for reliable impact-SE estimates, and three events (hospitalization, suicidal thoughts, spasms/tics) were not reported at all.

Table 4 describes the AE profile for specifically anxiety and chronic pain patients. While increased appetite, fatigue, and anxiety were the top three AEs overall (23.8%, 20.3%, 19.9%; Table 4), their within-subgroup proportions are uniformly lower, ranging 13–18% (Table 3), reflecting that no single symptom dominates any one condition. “Feeling sick or nauseous” and “increased tolerance” each account for ≈12% overall but varied by subgroup. Many low-frequency events in Table 3 (*<* 3% overall) appear only sporadically (*<* 3.5%) or not at all in subgroups presented in Table 4.

**Table 4:** Adverse Event Frequency by Subgroup: Anxiety vs. Chronic Pain.

| Adverse Event | Anxiety (n=108) | Chronic Pain (n=96) |
| --- | --- | --- |
| Increase in appetite | 15 (13.9%) | 17 (17.7%) |
| Tiredness/fatigue | 14 (13.0%) | 15 (15.6%) |
| Anxiety | 16 (14.8%) | 14 (14.6%) |
| Feeling sick or nauseous | 7 (6.5%) | 8 (8.3%) |
| Increased tolerance to cannabis medication | 10 (9.3%) | 5 (5.2%) |
| Paranoia | 11 (10.2%) | 3 (3.1%) |
| Memory loss | 6 (5.6%) | 3 (3.1%) |
| Weight gain | 4 (3.7%) | 6 (6.3%) |
| Dizziness | 2 (1.9%) | 4 (4.2%) |
| Other | 4 (3.7%) | 1 (1.0%) |
| Sore throat | 0 (0.0%) | 3 (3.1%) |
| Decreased appetite | 3 (2.8%) | 2 (2.1%) |
| Diarrhea | 1 (0.9%) | 3 (3.1%) |
| Loss of balance | 0 (0.0%) | 3 (3.1%) |
| Irritability | 2 (1.9%) | 1 (1.0%) |
| Hallucinations (visual/auditory) | 1 (0.9%) | 1 (1.0%) |
| Constipation | 2 (1.9%) | 0 (0.0%) |
| Negative behavioral/mood change | 1 (0.9%) | 2 (2.1%) |
| Somnolence | 1 (0.9%) | 0 (0.0%) |
| Loss of interest in usual activities | 1 (0.9%) | 2 (2.1%) |
| Agitation | 2 (1.9%) | 0 (0.0%) |
| Weakness | 2 (1.9%) | 0 (0.0%) |
| Increased pain sensitivity | 2 (1.9%) | 0 (0.0%) |
| Wheezing | 0 (0.0%) | 1 (1.0%) |
| Seizures | 0 (0.0%) | 0 (0.0%) |
| Increase in depression | 0 (0.0%) | 1 (1.0%) |
| Weight loss | 0 (0.0%) | 1 (1.0%) |
| Euphoria | 0 (0.0%) | 0 (0.0%) |
| Tremors | 1 (0.9%) | 0 (0.0%) |
| Hospitalization | 0 (0.0%) | 0 (0.0%) |
| Suicidal thoughts | 0 (0.0%) | 0 (0.0%) |
| Spasms/tics | 0 (0.0%) | 0 (0.0%) |
| **Total reports** | **108 (100%)** | **96 (100%)** |

**Note:** Adverse events reported for specifically anxiety and chronic pain.

In univariate logistic regression (Table 5), we examined the crude association between each candidate covariate and the odds of reporting any AE, separately within chronic pain and anxiety patients. Among the anxiety patients, having been to the doctor (OR 3.425, 95% CI 2.215–5.437), the number of unhealthy weeks (1-2 weeks vs. less than 1 week: OR 1.956, 95% CI 1.126, 3.435; 2-3 weeks vs. less than 1 week: OR 1.987, 95% CI 1.102, 3.584), routes of administration (Vape vs. dried flower: OR 1.74, 95% CI 1.039, 2.915), and product type (Equal ratio vs. High THC: OR 0.317, 95% CI 0.115, 0.947) were among the few predictors reaching statistical significance, while demographic and exposure variables remained largely nonsignificant. For chronic pain, age displayed a small but statistically significant predictor of the odds of having an AE, (age: OR 0.984, 95% CI 0.970, 0.999) as did the number of unhealthy weeks (2-3 weeks vs. less than 1 week: OR 1.663, 95% CI 1.003, 2.824).

**Table 5:**
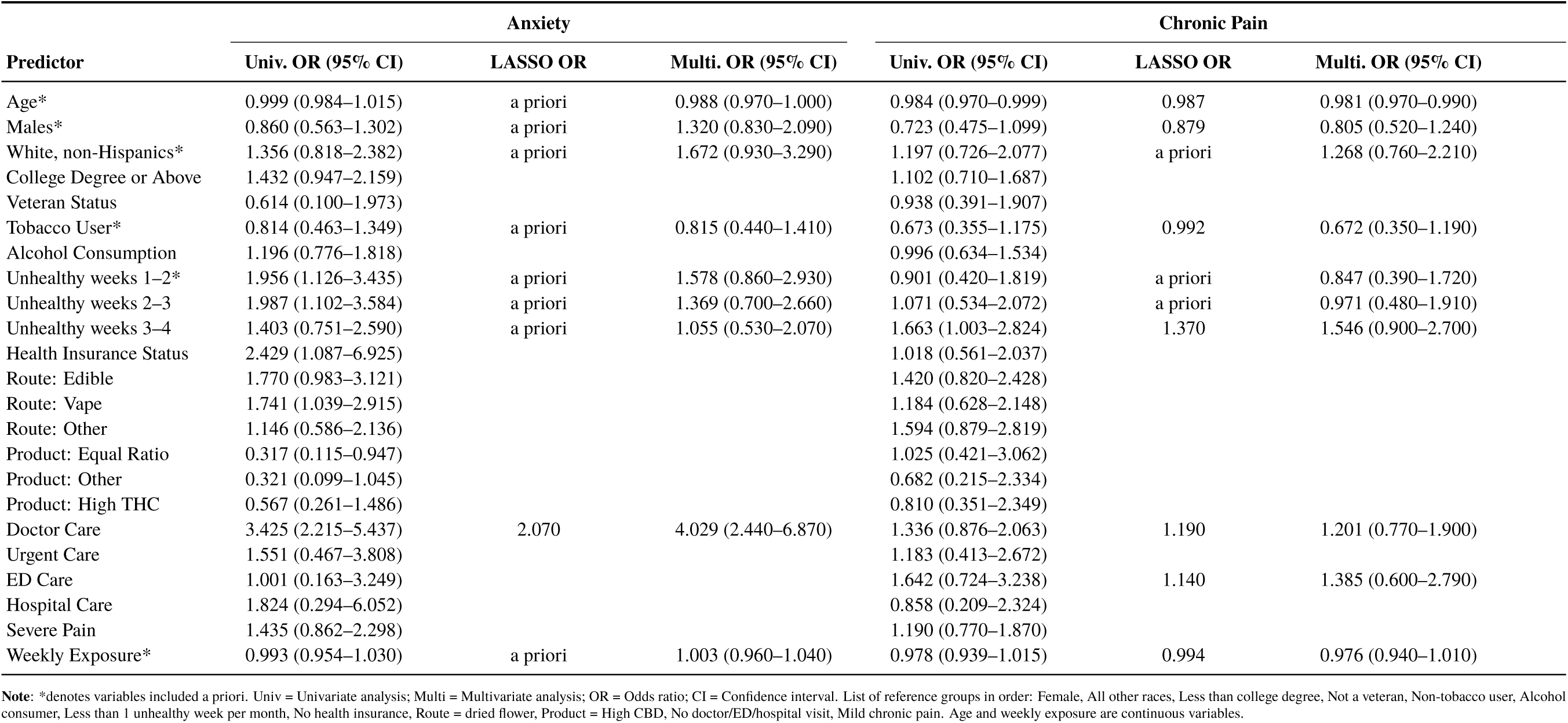
Univariate, LASSO, and Multivariate Logistic Regression for Adverse Event Odds Ratio.

To reduce model complexity and mitigate overfitting, we then applied LASSO penalization within each condition-specific dataset (Table 5). For anxiety patients, only visiting a doctor was a significant predictor of increased odds of having an AE. For chronic pain patients, age, sex, smoking status, having 3-4 unhealthy weeks per month, receiving doctor care, receiving ED care, and weekly exposure were significant predictors of AE Odds.

Our multivariate logistic models combined the a priori confounders (age, sex, race/ethnicity, smoking status, number of unhealthy weeks, and weekly cannabis exposure), and the LASSO-identified predictors (Table 5). Among those with anxiety, having visited the doctor was the only significant predictor of having an AE (OR 4.03, 95% CI 2.44–6.87); no other covariate was related to having an AE. For those with chronic pain, age was the only significant predictor of having an AE (OR 0.98, 95% CI 0.97–0.99).

The marginal exposure–response curves (Figure 1) plot the adjusted probability of reporting any AE against weekly cannabis use frequency separately for anxiety and chronic pain patients. The chronic pain curve shows a slight downward slope, AE probability increases from 3% to 2.1% across the exposure range, whereas the anxiety curve is virtually unchanged, hovering around 2% throughout. The narrow logit-scale 95% confidence bands indicate precision in these estimates. Overall, after adjusting for age, sex, race/ethnicity, smoking status, doctor visits, and unhealthy-weeks, weekly cannabis frequency does not appear to influence AE risk in either condition.

**Figure 1:**
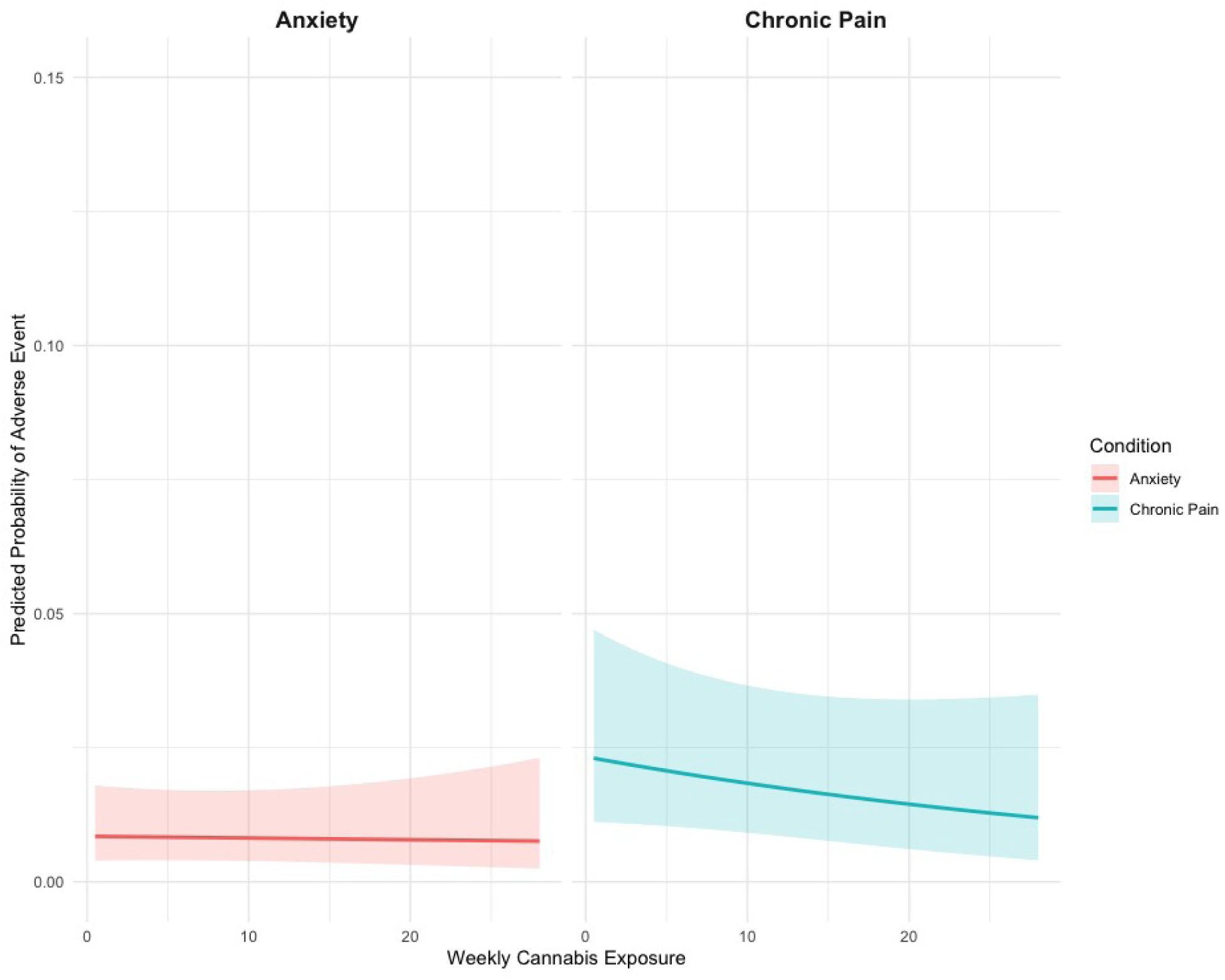
Marginal exposure-response curves stratified by anxiety and chronic pain patients.

Figure 2 presents the same marginal exposure–response curves stratified by key demographics: sex (Male vs. Female) and race/ethnicity (White non-Hispanic vs. all other race/ethnicities). Among all four plots, the slopes remain flat with increasing exposure: the maximal difference between strata is under one percentage point, and the bands overlap substantially. These plots confirm that sex and race produce small vertical shifts in baseline AE probability but do not modify the near-null exposure–response relationship.

**Figure 2:**
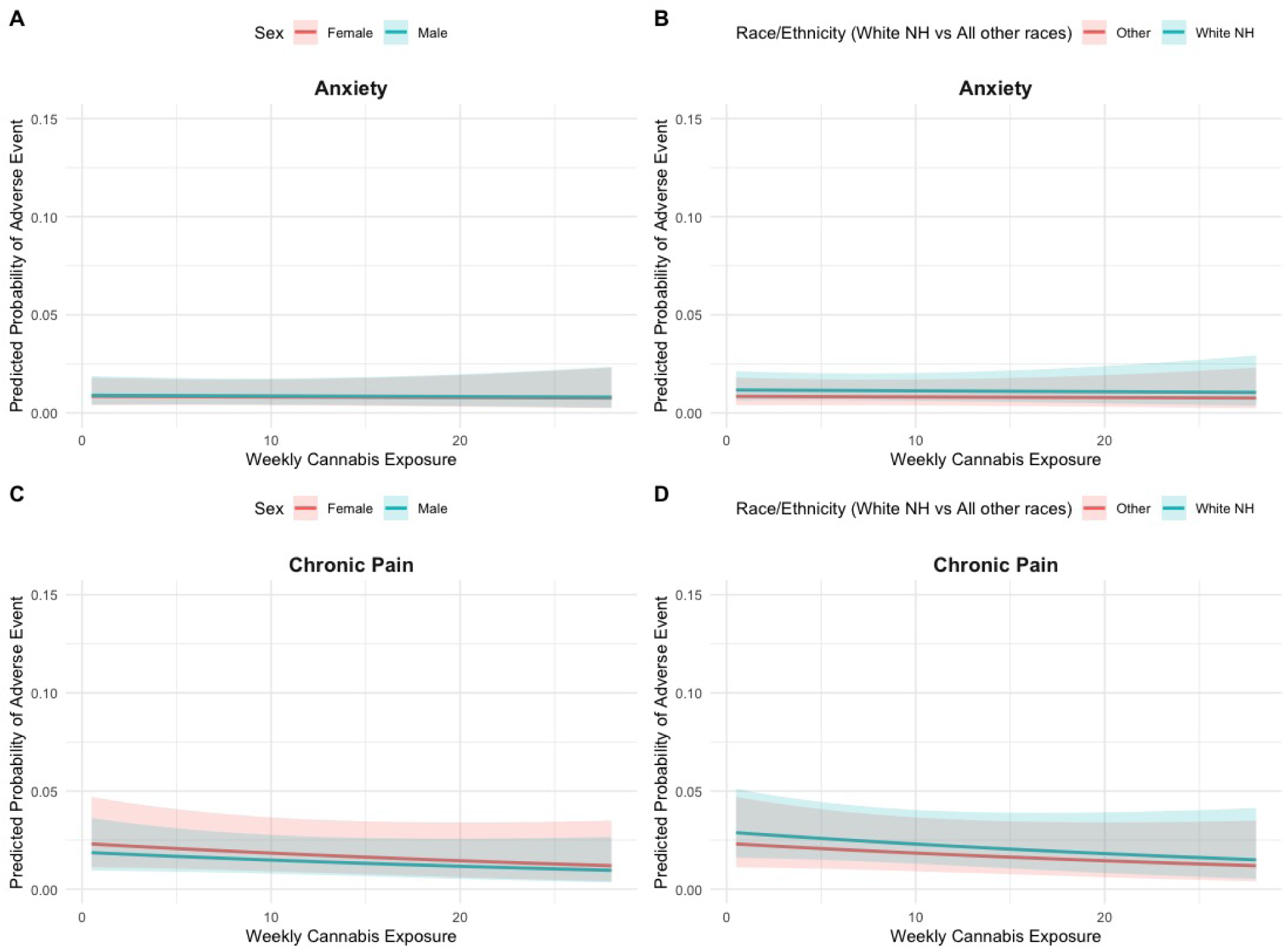
Marginal exposure-response curve across sex and race independent of medical condition.

Figure 3 depicts the adjusted probability of an AE as a function of patient age (with weekly exposure held at its mean) in the chronic pain and anxiety groups. Both curves slope gently downward: younger patients (age 20) have predicted AE probabilities of 3.0–3.2%, which decline to 2.0–2.2% by age 80. The confidence ribbons widen slightly at the extremes, reflecting fewer observations, but remain well within 1–5%. This indicates that, independent of cannabis dose and other confounders, older patients are slightly less likely to report AEs, possibly reflecting age-related reporting biases or differential susceptibility, agreeing with findings from our multivariate models.

**Figure 3:**
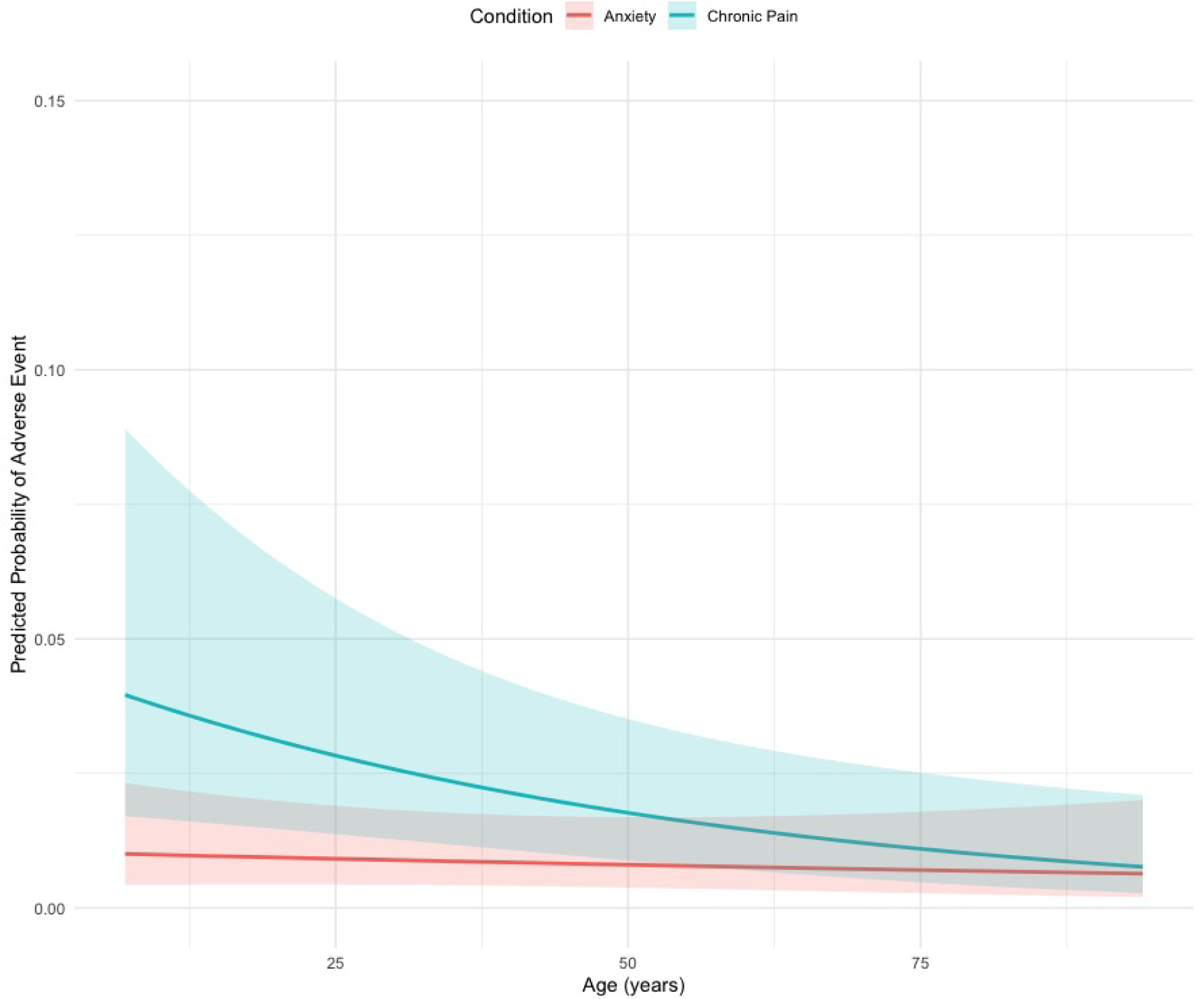
Marginal exposure-response curve across age independent of medical condition.

Given our 59.7 percent one-year follow-up rate, we conducted an ad hoc sensitivity analysis to assess how non–missing-at-random loss to follow-up might bias our reported 2.6 percent AE rate (see Figure 4). We assumed that the 40.3 percent of eligible patients who did not return for recertification could have experienced AEs at rates ranging from 5 percent (consistent with the lower end of registry estimates) up to 50 percent (upper estimates in small cohorts). For each assumed nonresponder AE percentage, we imputed the corresponding number of events among non-responders and recalculated the overall AE rate in the full cohort. Under a 5 percent assumption, the aggregate AE rate increases from 2.6% to approximately 3.55%, whereas a 50 percent assumption among all non-respondents yields an overall rate near 21.68%. This exploratory analysis illustrates that, if non-responders systematically experienced more side effects, our observed AE frequency likely represents a conservative lower bound.

**Figure 4:**
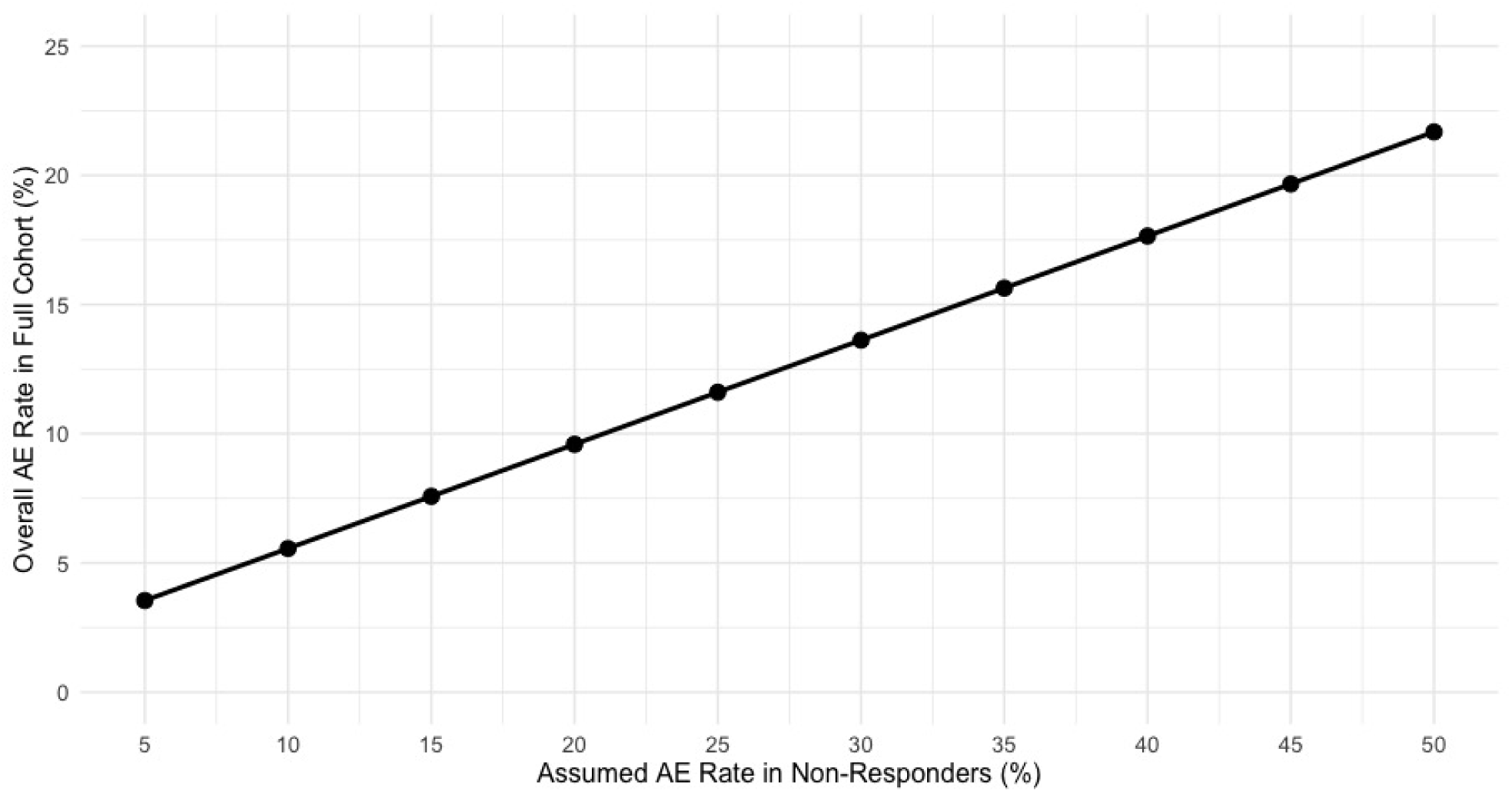
Sensitivity of overall adverse-event rate to non-responder risk assumptions.

## 4 Discussion

In our registry of more than 14,000 patients who used medical cannabis for one year, AEs proved rare and generally tolerable. Only 2.5 percent of users reported any AE, and when they did, the most common complaints, increased appetite (23.8 percent of AE reporters), fatigue (20.3 percent), and anxiety (19.9 percent), scored below 4 on a 0-to-10 impact scale (Table 3). Importantly, once we adjusted for patient characteristics and health status, weekly cannabis exposure was not associated with an increased odds of having an AE.

Notably, age proved to be a significant predictor of AE odds among those with chronic pain. With every 1 unit increase in age, the odds of having an AE reduced 1.6%. Among those with anxiety, the strongest predictor was having seen a doctor for one’s condition. Patients who had a doctor visit were about 4 times greater odds of reporting an AE in the anxiety subgroup. This finding likely suggests that patients who seek more medical care may have more complex health profiles. They may be on multiple medications, have more severe symptoms, or have comorbidities that heighten the likelihood of side effects once medical cannabis is added. From a clinical standpoint, providers should pay particular attention to co-medications and overall health statuses when initiating or adjusting cannabis treatment. Ensuring that patients understand potential interactions and monitoring them closely may be as important as dosing decisions.

Our results are consistent with randomized trials and international registries showing that medical cannabis is generally safe and well tolerated Pratt et al. (2019); Wang et al. (2008). Controlled trials of medical cannabis use have reported AE rates between 10 percent and 50 percent with almost exclusively mild events such as dizziness, dry mouth, and sedation and low discontinuation rates (Abrams et al., 2020; AminiLari et al., 2022; Aran et al., 2021; Grimison et al., 2024; Zylla et al., 2021). National cohorts in the UK and Canada similarly documented AE frequencies from 3 percent to 35 percent, again dominated by non-serious symptoms and without clear dose-response trends (Dalavaye et al., 2023; Ergisi et al., 2022; Erridge et al., 2022, 2023; Harris et al., 2022; Nicholas et al., 2023; Nimalan et al., 2022; Olsson et al., 2023; Pillai et al., 2022; Rifkin-Zybutz et al., 2023; Tait et al., 2023; Vivek et al., 2024). What our analysis adds is a large, US telehealth registry perspective with one year of follow-up data across multiple product types and routes of administration.

In the test of differences in log-odds trends, we did see meaningful differences in AE risk at lower weekly cannabis exposure (p-value = 0.036, See Table 2). However, our regression approach identified that the crude association between weekly cannabis exposure and AE odds were non-significant once we accounted for demographics, health-care utilization, and general health status. Taken together, the concordance of trial, registry, and real-world findings reinforces that, under clinical supervision, medical cannabis can be prescribed with a tolerable safety profile.

In our univariate analyses, specific routes of administration were associated with higher odds of reporting an AE among patients with anxiety. When we applied LASSO penalization within each condition subgroup, none of the route variables retained importance. This divergence suggests that although certain administration routes may appear to carry elevated risk on their own, their apparent effects are largely explained by confounding factors such as patients’ overall health status, frequency of doctor visits, or concurrent use habits. In other words, once we account for the broader clinical and demographic context, the choice of delivery method does not independently predict AEs. Clinicians should therefore consider focusing on patient complexity and co-medication profiles rather than route alone when evaluating safety. For future research exploring AE risk, careful adjustment for variables related to route of administration, such as product potency, dose per administration, and patient selection factors, is a necessary step to avoid biased estimates and to isolate any true effects of delivery method.

Given that we found an overall AE rate of *<*3%, independent of underlying condition, and that, in our multivariate models for both chronic-pain and anxiety patients, neither sex, race, smoking status, nor weekly cannabis exposure predicted AE risk, clinicians can counsel patients that the likelihood of experiencing a new side effect is likely low when cannabis is used as prescribed. Rather than focusing on dose adjustments alone, providers should prioritize comprehensive medication reviews and management of co-morbid health issues, particularly for patients with frequent healthcare utilization, since these factors more strongly signal AE reporting. In practice, this may include screening for poly pharmacy and potential drug–cannabis interactions before initiation, setting patient expectations that mild symptoms such as appetite changes or fatigue may occur but are generally tolerable, and scheduling follow-up visits not to adjust dose per se but to monitor overall clinical complexity and intervene in modifiable risk factors. By integrating these insights, clinicians can optimize safety while maintaining the therapeutic benefits of medical cannabis.

Our application of marginal exposure-response curves in a large sample of medical cannabis users is a novel contribution, providing clear, adjusted visualizations of how exposure relates to safety in real-world practice. However, it is important to recognize a key limitation of our exposure metric. We measured weekly exposure only by asking patients how many days per week they used cannabis and their regular serving size per use. This structure implicitly assumes one use session per day at whatever typical dose. We did not collect data on the number of times per day or the number of servings per session. Thus, a patient who uses cannabis four or five times daily would be treated the same as one who uses once per week, provided both fall into the same “every day” category. If we had also captured use per day, the true relationship between cumulative exposure and AE risk might differ. Future studies should therefore incorporate detailed frequency metrics to more accurately characterize dose–response and avoid underestimating risk among heavy, high-frequency users.

Although our cross-sectional design provides an important safety snapshot, longitudinal data is needed to understand incident AEs and their time course. Future work should link registry records with electronic health-record data to capture emergency department visits, hospitalizations, and serious events that may not be self-reported. Incorporating pharmacokinetic measures and detailed chemovar profiles, THC/CBD ratios, terpene content, formulation, would enable more precise modeling of dose-response relationships. Adaptive or pragmatic trial designs could randomize patients to different exposure levels with real-time safety monitoring, identifying thresholds beyond which AE risk increases. Because healthcare utilization emerged as a key signal, studies of drug-drug interactions in patients on polypharmacy regimens are also critical. Qualitative research on how patients perceive and report side effects could improve pharmacovigilance capture in real-world practice. Finally, expanding surveillance to other subgroups will ensure that safety conclusions apply broadly across all medical-cannabis users.

Our ad hoc sensitivity analysis reaffirms that, under missing–not–at–random scenarios, our observed 2.6 percent AE rate likely underestimates true risk but remains consistent with existing real-world pharmacovigilance data. When we impute a modest 5% AE rate among non-responders, the overall rate rises only to 3.6 percent, still below the 10–35 percent rates commonly reported in U.K. and Canadian registry studies, and even an extreme 50% assumption yields 21.7 percent, which falls squarely within the broad 3–57 percent range observed in smaller cohorts and meta-analyses (Crescioli et al., 2020; Ergisi et al., 2022; Erridge et al., 2023; Harris et al., 2022; Kalaba et al., 2022; Nicholas et al., 2023; Nimalan et al., 2022; Olsson et al., 2023; Rifkin-Zybutz et al., 2023; Tait et al., 2023). This suggests that our lower bound is plausible and that, while loss to follow-up may conceal some events, it is unlikely to shift the safety profile into the high-risk territory implied by controlled-trial discontinuation rates (typically *<*5 percent) or the serious-AE rates (*<*1–3 percent).

### 4.1 Strengths and limitations

Our study’s strengths include a large, national telehealth sample with minimal missing data and a 59% response rate, giving us the power to detect even modest safety signals. We combined univariate screening, cross-validated LASSO selection, and adjustment for a priori confounders (age, sex, race-ethnicity, smoking status, unhealthy weeks and weekly cannabis exposure) to build parsimonious, robust multivariate models. Nonetheless, because our data are cross-sectional and self-reported, we cannot establish temporality or causality, and recall bias may affect both exposure and event reporting. Our telehealth sample, while large, may not represent all medical cannabis patients, and we did not formally assess model discrimination or adjust for multiple comparisons beyond penalization. There are likely unmeasured factors that could also influence event reporting and are not included in our analysis. Finally, patient-rated impact scores lack clinical adjudication and may under capture rare but serious outcomes. Despite these limitations, our multi-method approach provides a rigorous framework for pharmacovigilance in medical cannabis therapy and informs both clinical practice and regulatory considerations.

### 4.2 Conclusion

Our large, real-world registry demonstrates that AEs from medical cannabis are infrequent, generally mild, and not driven by weekly cannabis exposure once complexity is accounted for. Healthcare engagement emerged as the key predictor of side-effect reporting for anxiety patients, highlighting the importance of comprehensive clinical assessment and medication review. These findings suggest that, with appropriate monitoring and attention to comorbidities and co-medications, medical cannabis can likely be prescribed safely. Future work using more granular exposure metrics and longitudinal designs will further refine our understanding of risk in diverse patient populations.

## Supporting information

Supplemental Material - Doucette et al., 2025

## Acknowledgements

Not Applicable.

## Author Contributions

Conceptualization, MLD, JC, EF; Methodology, MLD; Validation, MLD; Formal Analysis, MLD; Investigation, MLD, JC, EF; Data Curation, MLD; Writing – Original Draft Preparation, MLD; Writing – Review Editing, MLD, JC, EF; Visualization, MLD; Supervision, MLD; Project Administration, MLD. EF and JC.

## Institutional Review Board Statement

This study was approved by BRANY IRB (IRB00000080) on October 1, 2023.

## Data Availability Statement

The dataset presented in this article is not readily available because the data is proprietary to Leafwell and are not publicly available. All analyses were conducted by researchers with approved access under internal data use agreements.

## References

Abrams, D., Couey, P., Dixit, N., Sagi, V., Hagar, W., Vichinsky, E., and, et al. (2020). Effect of inhaled cannabis for pain in adults with sickle cell disease: A randomized clinical trial. JAMA Network Open, 3:e2010874.

AminiLari, M., Wang, L., Neumark, S., Adli, T., Couban, R., Giangregorio, A., and, et al. (2022). Medical cannabis and cannabinoids for impaired sleep: a systematic review and meta-analysis of randomized clinical trials. Sleep, 45:zsab234.

Aran, A., Harel, M., Cassuto, H., Polyansky, L., Schnapp, A., Wattad, N., and, et al. (2021). Cannabinoid treatment for autism: a proof-of-concept randomized trial. Molecular Autism, 12:6.

Armitage, P. (1955). Tests for linear trends in proportions and frequencies. Biometrics, 11(3):375–386.

Boehnke, K., Sinclair, R., Gordon, F., Hosanagar, A., Roehler, D., Smith, T., and, et al. (2024). Trends in u.s. medical cannabis registrations, authorizing clinicians, and reasons for use from 2020 to 2022. Annals of Internal Medicine, 177:458–466.

Cochran, W. G. (1954). Some methods for strengthening the common chi-squared tests. Biometrics, 10(4):417–451.

Cook, A., Sirmans, E., and Stype, A. (2023). Medical cannabis laws lower individual market health insurance premiums. International Journal of Drug Policy, 119:104143.

Crescioli, G., Lombardi, N., Bettiol, A., Menniti-Ippolito, F., Da Cas, R., Parrilli, M., and, et al. (2020). Adverse events following cannabis for medical use in tuscany: An analysis of the italian phytovigilance database. British Journal of Clinical Pharmacology, 86:106–120.

Dalavaye, N., Erridge, S., Nicholas, M., Pillai, M., Bapir, L., Holvey, C., and, et al. (2023). The effect of medical cannabis in inflammatory bowel disease: analysis from the uk medical cannabis registry. Expert Review of Gastroenterology & Hepatology, 17:85–98.

Doucette, M., Casarett, D., Hemraj, D., Grelotti, D., Macfarlan, D., and Fisher, E. (2024a). Towards a comprehensive understanding of medical conditions among medical cannabis patients in a large database: A descriptive analysis. Population Medicine. [Accessed 2024–11-06].

Doucette, M., Fisher, E., Hemraj, D., Kasabuski, M., and Chin, J. (2025a). Virtual care and medical cannabis access: A geospatial study of telemedicine’s role in reducing socioeconomic disparities. Telemedicine and e-Health.

Doucette, M., Hemraj, D., Bruce, D., Fisher, E., and Macfarlan, D. (2024b). Medical cannabis patients under the age of 21 in the united states: Description of demographics and conditions from a large patient database, 2019–2023. Adolescent Health, Medicine and Therapeutics, 15:63–72.

Doucette, M., Hemraj, D., Fisher, E., and Macfarlan, D. (2024c). Measuring the impact of medical cannabis law adoption on employer-sponsored health insurance costs: A difference-in-difference analysis, 2003–2022. Applied Health Economics and Health Policy.

Doucette, M., Kasabuski, M., Fisher, E., Chin, J., Bruce, D., and Kitsantas, P. (2025b). Identifying predictors of benzodiazepine discontinuation in medical cannabis patients with post-traumatic stress disorder using a machine learning approach. [Preprint; accessed 2025–03-07].

Doucette, M., Macfarlan, D., Kasabuski, M., Chin, J., and Fisher, E. (2024d). Impact of medical cannabis treatment on healthcare utilization in patients with post-traumatic stress disorder: A retrospective cohort study. [Preprint; accessed 2025–01-06].

Ergisi, M., Erridge, S., Harris, M., Kawka, M., Nimalan, D., Salazar, O., and, et al. (2022). Uk medical cannabis registry: an analysis of clinical outcomes of medicinal cannabis therapy for generalized anxiety disorder. Expert Review of Clinical Pharmacology, 15:487–495.

Erridge, S., Holvey, C., Coomber, R., Hoare, J., Khan, S., Platt, M., and, et al. (2023). Clinical outcome data of children treated with cannabis-based medicinal products for treatment resistant epilepsy—analysis from the uk medical cannabis registry. Neuropediatrics, 54:174–181.

Erridge, S., Kerr-Gaffney, J., Holvey, C., Coomber, R., Barros, D., Bhoskar, U., and, et al. (2022). Clinical outcome analysis of patients with autism spectrum disorder: analysis from the uk medical cannabis registry. Therapeutic Advances in Psychopharmacology, 12:20451253221116240.

Friedman, J., Hastie, T., and Tibshirani, R. (2008). glmnet: Lasso and Elastic-Net Regularized Generalized Linear Models. CRAN. R package version 1.0, published 2008-05-14.

Gottschling, S., Ayonrinde, O., Bhaskar, A., Blockman, M., D’Agnone, O., Schecter, D., and, et al. (2020). Safety considerations in cannabinoid-based medicine. International Journal of General Medicine, 13:1317–1333.

Greywoode, R., Cunningham, C., Hollins, M., and Aroniadis, O. (2023). Medical cannabis use patterns and adverse effects in inflammatory bowel disease. Journal of Clinical Gastroenterology, 57:824–829.

Grimison, P., Mersiades, A., Kirby, A., Tognela, A., Olver, I., Morton, R., and, et al. (2024). Oral cannabis extract for secondary prevention of chemotherapy-induced nausea and vomiting: Final results of a randomized, placebo-controlled, phase ii/iii trial. Journal of Clinical Oncology, 42:4040–4050.

Hachem, Y., Moride, Y., Castilloux, A., Castillon, G., Kalaba, M., Néron, A., and, et al. (2024). A descriptive analysis of adverse event reports from the quebec cannabis registry. Drug Safety, 47:161–171.

Hallinan, C., Gunn, J., and Bonomo, Y. (2022). Use of electronic medical records to monitor the safe and effective prescribing of medicinal cannabis: is it feasible? Australian Journal of Primary Health, 28:564–572.

Harris, M., Erridge, S., Ergisi, M., Nimalan, D., Kawka, M., Salazar, O., and, et al. (2022). Uk medical cannabis registry: an analysis of clinical outcomes of medicinal cannabis therapy for chronic pain conditions. Expert Review of Clinical Pharmacology, 15:473–485.

Hossain, M. and Chae, H. (2024). Medical cannabis: From research breakthroughs to shifting public perceptions and ensuring safe use. Integrative Medicine Research, 13:101094.

Jeddi, H., Busse, J., Sadeghirad, B., Levine, M., Zoratti, M., Wang, L., and, et al. (2024). Cannabis for medical use versus opioids for chronic non-cancer pain: a systematic review and network meta-analysis of randomised clinical trials. BMJ Open, 14:e068182.

Kalaba, M., Eglit, G., Feldner, M., Washer, P., Ernenwein, T., Vickery, A., and, et al. (2022). Longitudinal relationship between the introduction of medicinal cannabis and polypharmacy: An australian real-world evidence study. International Journal of Clinical Practice, 2022:8535207.

Krediet, E., Janssen, D., Heerdink, E., Egberts, T., and Vermetten, E. (2020). Experiences with medical cannabis in the treatment of veterans with ptsd: Results from a focus group discussion. European Neuropsychopharmacology, 36:244–254.

Lu, H. and Mackie, K. (2016). An introduction to the endogenous cannabinoid system. Biological Psychiatry, 79:516–525.

Lynskey, M., Athanasiou-Fragkouli, A., Thurgur, H., Schlag, A., and Nutt, D. (2024). Medicinal cannabis for treating post-traumatic stress disorder and comorbid depression: real-world evidence. BJPsych Open, 10:e62.

MacCallum, C., Lo, L., and Boivin, M. (2021). “is medical cannabis safe for my patients?” a practical review of cannabis safety considerations. European Journal of Internal Medicine, 89:10–18.

MacCallum, C. and Russo, E. (2018). Practical considerations in medical cannabis administration and dosing. European Journal of Internal Medicine, 49:12–19.

Mielenz, T., Jackson, E., Currey, S., DeVellis, R., and Callahan, L. F. (2006). Psychometric properties of the centers for disease control and prevention health-related quality of life (cdc hrqol) items in adults with arthritis. Health and Quality of Life Outcomes, 4:66.

Nacasch, N., Avni, C., and Toren, P. (2023). Medical cannabis for treatment-resistant combat ptsd. [Accessed 2024–02-14].

National Academies of Sciences, Engineering, and Medicine (2017). The health effects of cannabis and cannabinoids: The current state of evidence and recommendations for research. [Accessed 2023-08-16].

Nguyen, T., Li, Y., Greene, D., Stancliff, S., and Quackenbush, N. (2023). Changes in prescribed opioid dosages among patients receiving medical cannabis for chronic pain, new york state, 2017–2019. JAMA Network Open, 6:e2254573.

Nicholas, M., Erridge, S., Bapir, L., Pillai, M., Dalavaye, N., Holvey, C., and, et al. (2023). Uk medical cannabis registry: assessment of clinical outcomes in patients with headache disorders. Expert Review of Neurotherapeutics, 23:85–96.

Nimalan, D., Kawka, M., Erridge, S., Ergisi, M., Harris, M., Salazar, O., and, et al. (2022). Uk medical cannabis registry palliative care patients cohort: initial experience and outcomes. Journal of Cannabis Research, 4:1–4.

Olsson, F., Erridge, S., Tait, J., Holvey, C., Coomber, R., Beri, S., and, et al. (2023). An observational study of safety and clinical outcome measures across patient groups in the united kingdom medical cannabis registry. Expert Review of Clinical Pharmacology, 16:257–266.

O’Connell, M., Sandgren, M., Frantzen, L., Bower, E., and Erickson, B. (2019). Medical cannabis: Effects on opioid and benzodiazepine requirements for pain control. Annals of Pharmacotherapy, 53:1081–1086.

Pillai, M., Erridge, S., Bapir, L., Nicholas, M., Dalavaye, N., Holvey, C., and, et al. (2022). Assessment of clinical outcomes in patients with post-traumatic stress disorder: analysis from the uk medical cannabis registry. Expert Review of Neurotherapeutics, 22:1009–1018.

Pratt, M., Stevens, A., Thuku, M., Butler, C., Skidmore, B., Wieland, L., and, et al. (2019). Benefits and harms of medical cannabis: a scoping review of systematic reviews. Systematic Reviews, 8:320.

Purcell, C., Davis, A., Moolman, N., and Taylor, S. (2019). Reduction of benzodiazepine use in patients prescribed medical cannabis. Cannabis and Cannabinoid Research, 4:214–218.

Radwan, M., Chandra, S., Gul, S., and ElSohly, M. (2021). Cannabinoids, phenolics, terpenes and alkaloids of cannabis. Molecules, 26:2774.

Rehman, Y., Saini, A., Huang, S., Sood, E., Gill, R., and Yanikomeroglu, S. (2021). Cannabis in the management of ptsd: a systematic review. AIMS Neuroscience, 8:414–434.

Ried, K., Tamanna, T., Matthews, S., and Sali, A. (2023). Medicinal cannabis improves sleep in adults with insomnia: a randomised double-blind placebo-controlled crossover study. Journal of Sleep Research, 32:e13793.

Rifkin-Zybutz, R., Erridge, S., Holvey, C., Coomber, R., Gaffney, J., Lawn, W., and, et al. (2023). Clinical outcome data of anxiety patients treated with cannabis-based medicinal products in the united kingdom: a cohort study from the uk medical cannabis registry. Psychopharmacology, 240:1735–1745.

Roitman, P., Mechoulam, R., Cooper-Kazaz, R., and Shalev, A. (2014). Preliminary, open-label, pilot study of add-on oral 9-tetrahydrocannabinol in chronic post-traumatic stress disorder. Clinical Drug Investigation, 34:587–591.

RStudio Team (2025). RStudio: Integrated Development Environment for R. RStudio, PBC, Boston, MA.

Schlag, A. (2020). An evaluation of regulatory regimes of medical cannabis: What lessons can be learned for the uk? Medical Cannabis and Cannabinoids, 3:76–83.

Shi, Y. (2017). Medical marijuana policies and hospitalizations related to marijuana and opioid pain reliever*. Drug and Alcohol Dependence, 173:144–150.

Shover, C. and Humphreys, K. (2019). Six policy lessons relevant to cannabis legalization. American Journal of Drug and Alcohol Abuse, 45:698–706.

Signorell, A., Aho, K., and Alfons, A, e. a. (2025). DescTools: Tools for Descriptive Statistics. CRAN. R package version 0.99.60, published 2025-03-28.

Sobotka, L., Mumtaz, K., Hinton, A., Kelly, S., Conteh, L., Michaels, A., and, et al. (2021). Cannabis use may reduce healthcare utilization and improve hospital outcomes in patients with cirrhosis. Annals of Hepatology, 23:100280.

Solmi, M., Toffol, M., Kim, J., Choi, M., Stubbs, B., Thompson, T., and, et al. (2023). Balancing risks and benefits of cannabis use: umbrella review of meta-analyses of randomised controlled trials and observational studies. BMJ, 382:e072348.

Sunil, M., Karimi, P., Leong, R., Zuniga-Villanueva, G., and Ratcliffe, E. (2022). Therapeutic effects of medicinal cannabinoids on the gastrointestinal system in pediatric patients: A systematic review. Cannabis and Cannabinoid Research, 7:769–776.

Sznitman, S., Vulfsons, S., Meiri, D., and Weinstein, G. (2020). Medical cannabis and insomnia in older adults with chronic pain: a cross-sectional study. BMJ Supportive & Palliative Care, 10:415–420.

Tait, J., Erridge, S., and Sodergren, M. (2023). Uk medical cannabis registry: A patient evaluation. Journal of Pain & Palliative Care Pharmacotherapy, 37:170–177.

Takakuwa, K., Hergenrather, J., Shofer, F., and Schears, R. (2020). The impact of medical cannabis on intermittent and chronic opioid users with back pain: How cannabis diminished prescription opioid usage. Cannabis and Cannabinoid Research, 5:263–270.

Tibshirani, R. (1996). Regression shrinkage and selection via the lasso. Journal of the Royal Statistical Society: Series B (Methodological), 58(1):267–288.

Tibshirani, R., Bien, J., Friedman, J., Hastie, T., Simon, N., Taylor, J., and Tibshirani, R. J. (2010). Strong rules for discarding predictors in lasso-type problems. arXiv preprint arXiv:1011.2234.

Vivek, K., Karagozlu, Z., Erridge, S., Holvey, C., Coomber, R., Rucker, J., and, et al. (2024). Uk medical cannabis registry: Assessment of clinical outcomes in patients with insomnia. Brain and Behavior, 14:e3410.

Von Korff, M., DeBar, L. L., Krebs, E. E., Kerns, R. D., Deyo, R. A., and Keefe, F. J. (2020). Graded chronic pain scale revised: Mild, bothersome, and high impact chronic pain. Pain, 161:651–661.

Walsh, J., Maddison, K., Rankin, T., Murray, K., McArdle, N., Ree, M., and, et al. (2021). Treating insomnia symptoms with medicinal cannabis: a randomized, crossover trial of the efficacy of a cannabinoid medicine compared with placebo. Sleep, 44:zsab149.

Wang, R., Bonomo, Y., and Hallinan, C. (2024). Overview of global monitoring systems for the side effects and adverse events associated with medicinal cannabis use: a scoping review using a systematic approach. BMJ Open, 14:e085166.

Wang, T., Collet, J., Shapiro, S., and Ware, M. (2008). Adverse effects of medical cannabinoids: a systematic review. Canadian Medical Association Journal, 178:1669–1678.

Wen, H. and Hockenberry, J. (2018). Association of medical and adult-use marijuana laws with opioid prescribing for medicaid enrollees. JAMA Internal Medicine, 178:673–679.

Zahran, H. S., Kobau, R., Moriarty, D. G., Zack, M. M., Holt, J., Donehoo, R., for Disease Control, C., and (CDC), P. (2005). Health-related quality of life surveillance—united states, 1993–2002. Morbidity and Mortality Weekly Report Surveillance Summaries, 54:1–35.

Zylla, D., Eklund, J., Gilmore, G., Gavenda, A., Guggisberg, J., VazquezBenitez, G., and, et al. (2021). A randomized trial of medical cannabis in patients with stage iv cancers to assess feasibility, dose requirements, impact on pain and opioid use, safety, and overall patient satisfaction. Supportive Care in Cancer, 29:7471–7478.

