## Supplemental Material - Doucette et al., 2025 for "Weed Out the Risk: Pharmacovigilance in Medical Cannabis Users"

**Supplemental Table 1:** Mapping of serving-size responses to standardized consumption units

| **Route of Administration** | **Response Option** | **Intensity (common units)** |
| --- | --- | --- |
| **Flower (Smoked)** | Less than 1 bowl or 1 joint | 0.5 |
|  | 1 bowl or 1 joint | 1 |
|  | 2 bowls or 2 joints | 2 |
|  | 3 or more bowls or joints | 3 |
| **Vape Pen / Inhaler** | 1 puff | 0.5 |
|  | 2 puffs | 1 |
|  | 3 puffs | 1.5 |
|  | 4 puffs | 2 |
|  | 5 or more puffs | 3 |
| **Tincture / Oil / Drops** | Less than 0.25 ml | 0.5 |
|  | 0.25 – 0.5 ml | 1 |
|  | 0.51 – 0.75 ml | 1.5 |
|  | 0.76 – 1.0 ml | 2 |
|  | 1 – 1.5 ml | 2.5 |
|  | 1.51 – 2 ml | 3 |
|  | Over 2 ml | 4 |
| **Sublingual Spray** | 1 spray | 0.5 |
|  | 2 sprays | 1 |
|  | 3 sprays | 1.5 |
|  | 4 sprays | 2 |
|  | 5 or more sprays | 3 |
| **Edibles / Tablets / Topical / Other** | Less than one serving (e.g., cutting an edible in half) | 0.5 |
|  | 1 serving (e.g., 1 edible, 1 tablet, 1 application) | 1 |
|  | 2 servings | 2 |
|  | 3 or more servings | 3 |
| **Note.** Intensities were multiplied by the self-reported number of days used per week to derive each participant’s weekly exposure. | | |
